# Real-world Effectiveness of BNT162b2 Against Infection and Severe Diseases in Children and Adolescents

**DOI:** 10.1101/2023.06.16.23291515

**Authors:** Qiong Wu, Jiayi Tong, Bingyu Zhang, Dazheng Zhang, Jiajie Chen, Yuqing Lei, Yiwen Lu, Yudong Wang, Lu Li, Yishan Shen, Jie Xu, L. Charles Bailey, Jiang Bian, Dimitri A. Christakis, Megan L. Fitzgerald, Kathryn Hirabayashi, Ravi Jhaveri, Alka Khaitan, Tianchen Lyu, Suchitra Rao, Hanieh Razzaghi, Hayden T. Schwenk, Fei Wang, Margot I. Witvliet, Eric J. Tchetgen Tchetgen, Jeffrey S. Morris, Christopher B. Forrest, Yong Chen

**Affiliations:** Department of Biostatistics, Epidemiology, and Informatics, University of Pennsylvania Perelman School of Medicine, Philadelphia, PA, USA; Department of Health Outcomes Biomedical Informatics, University of Florida, Gainesville, FL, USA; Applied Clinical Research Center, The Children’s Hospital of Philadelphia, Philadelphia, PA, USA; Center for Child Health, Behavior, and Development, Seattle Children’s Research Institute, Seattle, WA, USA; Department of Medicine, Grossman School of Medicine, New York University, New York, NY, USA; Division of Pediatric Infectious Diseases, Ann & Robert H. Lurie Children’s Hospital of Chicago, Chicago, IL, USA; Department of Pediatrics, Ryan White Center for Pediatric Infectious Diseases and Global Health, Indiana University School of Medicine, IN, USA; Department of Pediatrics, University of Colorado School of Medicine and Children’s Hospital Colorado, Aurora, CO, USA; Department of Pediatrics, Stanford School of Medicine, Stanford, CA, USA; Department of Population Health Sciences, Weill Cornell Medicine, New York, NY, USA; Department of Sociology, Social Work and Criminal Justice, Lamar University, Beaumont, TX, USA; Department of Statistics and Data Science, The Wharton School, The University of Pennsylvania, PA, USA

## Abstract

**Background:** The efficacy of the BNT162b2 vaccine in pediatrics was assessed by randomized trials before the Omicron variant’s emergence. The long-term durability of vaccine protection in this population during the Omicron period remains limited.

**Objective:** To assess the effectiveness of BNT162b2 in preventing infection and severe diseases with various strains of the SARS-CoV-2 virus in previously uninfected children and adolescents.

**Design:** Comparative effectiveness research accounting for underreported vaccination in three study cohorts: adolescents (12 to 20 years) during the Delta phase, children (5 to 11 years) and adolescents (12 to 20 years) during the Omicron phase.

**Setting:** A national collaboration of pediatric health systems (PEDSnet).

**Participants:** 77,392 adolescents (45,007 vaccinated) in the Delta phase, 111,539 children (50,398 vaccinated) and 56,080 adolescents (21,180 vaccinated) in the Omicron period.

**Exposures:** First dose of the BNT162b2 vaccine vs. no receipt of COVID-19 vaccine.

**Measurements:** Outcomes of interest include documented infection, COVID-19 illness severity, admission to an intensive care unit (ICU), and cardiac complications. The effectiveness was reported as (1-relative risk)*100% with confounders balanced via propensity score stratification.

**Results:** During the Delta period, the estimated effectiveness of BNT162b2 vaccine was 98.4% (95% CI, 98.1 to 98.7) against documented infection among adolescents, with no significant waning after receipt of the first dose. An analysis of cardiac complications did not find an increased risk after vaccination. During the Omicron period, the effectiveness against documented infection among children was estimated to be 74.3% (95% CI, 72.2 to 76.2). Higher levels of effectiveness were observed against moderate or severe COVID-19 (75.5%, 95% CI, 69.0 to 81.0) and ICU admission with COVID-19 (84.9%, 95% CI, 64.8 to 93.5). Among adolescents, the effectiveness against documented Omicron infection was 85.5% (95% CI, 83.8 to 87.1), with 84.8% (95% CI, 77.3 to 89.9) against moderate or severe COVID-19, and 91.5% (95% CI, 69.5 to 97.6)) against ICU admission with COVID-19. The effectiveness of the BNT162b2 vaccine against the Omicron variant declined after 4 months following the first dose and then stabilized. The analysis revealed a lower risk of cardiac complications in the vaccinated group during the Omicron variant period.

**Limitations:** Observational study design and potentially undocumented infection.

**Conclusions:** Our study suggests that BNT162b2 was effective for various COVID-19-related outcomes in children and adolescents during the Delta and Omicron periods, and there is some evidence of waning effectiveness over time.

**Primary Funding Source:** National Institutes of Health

## INTRODUCTION

The Food and Drug Administration (FDA) expanded the emergency use authorization of the BNT162b2 mRNA COVID-19 vaccine (Pfizer-BioNTech) to 12-15-year-olds on May 10, 2021, and to 5-11-year-olds on October 29, 2021. As of April 5, 2023, the Centers for Disease Control and Prevention (CDC) reports indicate that 40% of U.S. children aged 5-to-11-year-olds and 72% of adolescents aged 12-to-18-year-olds had received at least one dose of the vaccine. The emergence of the Omicron variant (B.1.1.529) and its subvariants in early 2022 led to a new surge in COVID-19 cases worldwide (1). The randomized trials of the BNT162b2 vaccine which demonstrated high efficacy of 2 doses against COVID-19 (100% and 91% among those aged 12-15 and 5-11 years, respectively) were conducted before the emergence of the Omicron variant (2,3).

Several observational studies have been conducted to investigate the effectiveness of vaccination in real-world settings (4–8). However, prior studies have had limited follow-up periods, covering the Delta variant or earlier subvariants of Omicron periods only. Studies evaluating the Omicron variant have only assessed the short-term effects of the vaccine, with only one study involving children evaluating the effect beyond 3 months (9). There is limited information on the long-term durability of vaccine protection during the Omicron period. Few existing studies on U.S. pediatric populations have covered both hospitalized patients and those with mild or asymptomatic conditions. Furthermore, while studies have acknowledged limitations due to misclassification in vaccination status in real-world effectiveness studies, none have rigorously evaluated the impacts of such misclassification nor accounted for the potential bias it may introduce.

To address these gaps in our knowledge of the pediatric effectiveness of SARS-CoV-2 vaccination, we designed this study to assess the real-world effectiveness of BNT162b2 among children and adolescents during the Delta and Omicron variant-predominant periods using electronic health record (EHR) data from a national network of U.S. pediatric medical centers. Our study used a novel comparative effectiveness research (CER) method and adjusted for underreporting issues in vaccination status and has several attractive features that strengthen reliability of our inference. First, it is the largest study to date in the U.S. estimating vaccine effectiveness in children and adolescents, covering a broad spectrum of the U.S. pediatric population. Second, the study examined the effectiveness against infection over a longer follow-up period than any previous study, enabling evaluation of the durability of vaccine protection. Third, the study included a diverse representation of U.S. pediatric populations from primary care, specialty care, emergency department, testing centers, and inpatient settings. Fourth, the study was the first to account for the incomplete capture of vaccination status by health systems in the U.S. Finally, in addition to infection and severe disease endpoints, we also studied the effect of vaccination on the incidence of myocarditis, pericarditis or multisystem inflammatory syndrome (MIS) to assess the effect of vaccination relative to potential risk of cardiac complications.

## METHODS

### DATA SOURCES

This study used EHR data from PEDSnet (10), which is a national collaboration of pediatric health systems that share EHR data, conduct research, and improve outcomes together. Participating institutions in this study included: Children’s Hospital of Philadelphia, Cincinnati Children’s Hospital Medical Center, Children’s Hospital Colorado, Ann & Robert H. Lurie Children’s Hospital of Chicago, Nationwide Children’s Hospital, Nemours Children’s Health System (inclusive of the Delaware and Florida health system), Seattle Children’s Hospital, and Stanford Children’s Health. Data were extracted from the PEDSnet COVID-19 Database Version Week 141 (11). A detailed description of EHR data is available in Section S1 of the Supplementary Appendix.

### SPECIFICATION OF HYPOTHETICAL TRIALS AND CER STUDIES

Hypothetical randomized controlled trials (RCT) were specified to guide the design of observational studies to assess the real-world effectiveness of treatments (12). We designed and conducted CER studies to investigate the effectiveness of the BNT162b2 vaccine in preventing infection with various strains of the SARS-CoV-2 virus in children and adolescents in the United States. The three study cohorts focused on documented SARS-CoV-2 infection and outcomes in:

- ***Study cohort 1*** (Delta study in adolescents): adolescents aged 12 to 20 years during the period when the Delta variant was prevalent from July 1, 2021, to November 30, 2021.
- ***Study cohort 2*** (Omicron study in children): children aged 5 to 11 years during the period when the Omicron variant was prevalent from January 1, 2022, to November 30, 2022.
- ***Study cohort 3*** (Omicron study in adolescents): adolescents aged 12 to 20 years during the period when the Omicron variant was prevalent from January 1, 2022, to November 30, 2022.

The design of hypothetical trials and implementation procedures in real-world data are summarized in Table S1 of the Supplementary Appendix.

Eligibility criteria included age of 5 to 11 years for children or 12 to 20 years for adolescents at the start of the study period and no previous COVID-19 vaccination or documented SARS-CoV-2 infection. Additionally, participants were required to have a prior encounter (including telephone or telehealth encounters) within 18 months of cohort entrance to ensure that they had an ongoing interaction with the health system.

The intervention of interest was vaccination, in comparison with no receipt of any type of COVID-19 vaccine. Since the BNT162b2 vaccine covered more than 85% of documented vaccinations among children and adolescents in the PEDSnet database, in this study we focused primarily on studying the effectiveness of this vaccine, although the supplementary appendix reports a sensitivity analysis investigating all types of reported COVID-19 assessed in the U.S., with 85.7% BNT162b2, 1.9% mRNA-1273, and 12.3% unspecified COVID-19 vaccine.

In this CER study using real-world data, the cohort entrance date for the intervention group was defined as the date of the first dose of the BNT162b2 vaccine, while for the comparator group, it was a randomly selected date from visits, chosen to ensure the distribution of index dates for the control group matched the distribution of index dates for the vaccination group to control for time effects. The risk period for the study began 28 days after the index date such that infections within 28 days were excluded.

Randomized trials achieve balance across potential confounders by randomly allocating the treatment to intervention and comparator groups. In our study, we attempted to balance the intervention and comparator groups by adjusting for a large number of measured confounders collected prior to cohort entry using propensity score stratification (13). We built the propensity score model based on demographic factors including age, sex, race/ethnicity, clinical factors including obesity status, a baseline chronic condition indicator as defined by the Pediatric Medical Complexity Algorithm (PMCA) (14), and a list of pre-existing chronic conditions, and healthcare utilization factors collected prior to the cohort entry including the number of inpatients, outpatients, ED visits, unique mediations, and the number of negative COVID-19 tests. We stratified the patients into propensity score quintiles based on these factors. See Table S3 in the Supplementary Appendix for detailed definitions of study variables.

The four COVID-19 outcomes of interest were: documented SARS-CoV-2 infection, mild COVID-19, moderate/severe COVID-19, and ICU admission with COVID-19. We did not evaluate death from COVID-19 as it was too rare among children and adolescents to study quantitatively. In our study, SARS-CoV-2 infections were defined by and occurrence of positive polymerase-chain-reaction (PCR), serology, or antigen tests or diagnoses of COVID-19, post-acute sequelae of SARS-CoV-2 (PASC), or multisystem inflammatory syndrome in children (MIS-C) regardless of the presence of symptoms. Classification of mild, moderate, or severe COVID-19 infections was defined based on the symptoms and health conditions diagnosed from 7 days prior to 13 days post the date of a documented COVID-19 infection as in Forrest et al. (2022) (15). ICU admission with COVID-19 was defined by any ICU visit 7 days prior to 13 days after documented SARS-CoV-2 infection. Additionally, we considered the clinical outcomes of cardiac complications identified as incidence of myocarditis, pericarditis or MIS to allow for a comprehensive capture of potential cardiac complications after infection and evaluate the effect of the BNT162b2 vaccine in terms of cardiac risks.

### STATISTICAL ANALYSIS

We evaluated covariate balancing after propensity score stratification by plotting the standardized mean differences (SMD) between variable values for the vaccinated and unvaccinated groups, with a difference of 0.1 or less indicating an acceptable balance. We used Poisson regression to estimate the relative risk between two treatment arms for the risk of each outcome while adjusting for different follow-up lengths among participants. Since immunization records are often captured and stored across multiple disconnected sources, resulting in incomplete vaccination records in patients’ EHRs, we mitigated the potential bias arising from this underreporting issue by incorporating an integration likelihood(16) of the Poisson regression with a pre-specified range of misclassification rates. The vaccine effectiveness was defined as 100*(1-relative risk). The details of statistical methods are described in Section S2 of the Supplementary Appendix.

We conducted secondary analyses stratified by 2-month intervals since receipt of vaccination to investigate the durability of vaccine protection. Subgroup analyses were also performed to investigate differences in vaccine protection according to age groups (5-to-8, 9-to-11, 12-to-15, 16-to-20).

### SENSITIVITY ANALYSES

Extensive sensitivity analyses were conducted to evaluate the robustness of the research findings; see Supplementary Appendix Sections S6 – S15, S19 for the impacts of cohort design. In scenarios in which any categorical covariates were unbalanced (with a standardized mean difference>0.1), we included a sensitivity analysis excluding participants in that category. A sensitivity analysis defining the risk period with no waiting period after the index date was conducted. To assess whether population heterogeneity, established by applying eligibility criteria that required a prior encounter within 18 months of cohort entry, could influence our vaccine effectiveness estimates, we conducted sensitivity analyses within a more restricted time window. A sensitivity analysis was conducted to compare the proposed method to a sequential target trial emulation pipeline not accounting for underreporting issues to assess the robustness of findings. Since the proportion of patients entered with ED visits is relatively low in the vaccinated cohort compared to the unvaccinated, we conducted a sensitivity analysis excluding all participants who entered the cohort during an ED visit. Residual study bias from unmeasured and systematic sources can still exist in observational studies after controlling for measured confounders; thus, we conducted negative control outcome experiments (13,17,18) in which the null hypothesis of no effect was believed to be true using 40 negative control outcomes pre-specified by pediatric physicians. The empirical null distribution and calibrated effectiveness were reported as sensitivity analyses. The relative risks for cardiac complications defined by myocarditis or pericarditis (excluding MIS) were estimated. We also reported the estimated vaccine effectiveness from all brands of COVID-19 vaccines. Given the prolonged presence of the Omicron variant and the emergence of several sub-variants, we conducted a secondary analysis to assess the vaccine effectiveness related to sub-variants of Omicron.

### MISSINGNESS IN VACCINE RECORDS

Vaccine status may be missing for individuals whose vaccine doses were administered by a site outside of the PEDSnet network care delivery sites. It is likely that patients recorded as vaccinated in the EHR are true positives, so specificity could be very high, but sensitivity would be reduced by undocumented vaccinations (false negatives). To account for potential bias from the underreporting issue in vaccination status, a range of possible sensitivities based on our prior study was pre-specified for each study. The sensitivity range was considered to be 0.8 to 1 for the study involving children and 0.7 to 0.9 for the studies involving adolescents. By accounting for the underreporting and specifying a range of sensitivity, the study aimed to minimize the impact of bias caused by the underreporting in the estimation of the effectiveness of the COVID-19 vaccine among children and adolescents. The details of statistical methods are described in Section S2 of the Supplementary Appendix.

To further evaluate the robustness of statistical methods we used to account for underreporting, we varied our CER method by considering alternative methods for bias correction, including the naive method (without adjusting for underreporting), using different ranges of misclassification rates, and using a fully Bayesian method (19). To evaluate the impact of differential misclassification on effectiveness estimates, we conducted sensitivity analyses simulating vaccination status according to various differential misclassification scenarios. Results from these sensitivity analyses are summarized in Sections S16-18 of the Supplementary Appendix.

## RESULTS

### STUDY POPULATION

A total of 77,392 adolescents (45,007 vaccinated) within the PEDSnet network were identified to study the effectiveness of vaccination against Delta infection and severe outcomes (see Table 1 for baseline characteristics). 111,539 children (50,398 vaccinated) and 56,080 adolescents (21,180 vaccinated) were included in the cohort to study the effectiveness of vaccination against the Omicron infections (see Table 2 for baseline characteristics). The vaccinated and unvaccinated groups had a slightly unbalanced distribution of testing rates before cohort entry across all three cohorts. After propensity-score stratification, all covariates were well balanced between vaccinated and unvaccinated groups with an SMD smaller than 0.1 in the Omicron study involving children (Figure S13) and involving adolescents (Figure S14). In the study evaluating vaccine effectiveness for adolescents during the Delta period, one site remained unbalanced after propensity-score stratification with an SMD larger than 0.1, and thus a sensitivity analysis was conducted by excluding participants from this site which gave consistent results with the primary analysis (see Figure S12 and Section S6 in Supplementary Appendix). Additional characteristics of study cohorts including additional medical conditions, deaths, follow-up durations were summarized in Section S5 of Supplementary Appendix.

**Table 1.** Baseline characteristics of adolescents 12 to 20 years of age in the study of the effectiveness of the BNT162b2 vaccine in preventing infection and severe diseases with SARS-CoV-2 during the period when the Delta variant was prevalent.

| *Delta study in adolescents* |  |  |  |
| --- | --- | --- | --- |
|  | **Vaccinated**<br>**(N=45,007)** | **Unvaccinated**<br>**(N=32,385)** | **Overall**<br>**(N=77,392)** |
| **Age** |  |  |  |
| Median [Q1, Q3] | 14 [13, 16] | 15 [13, 17] | 15 [13, 16] |
| **Distribution** |  |  |  |
| 12 | 8922 (19.8%) | 4926 (15.2%) | 13848 (17.9%) |
| 13 | 8099 (18.0%) | 4864 (15.0%) | 12963 (16.7%) |
| 14 | 8037 (17.9%) | 4923 (15.2%) | 12960 (16.7%) |
| 15 | 7311 (16.2%) | 4749 (14.7%) | 12060 (15.6%) |
| 16 | 5450 (12.1%) | 4546 (14.0%) | 9996 (12.9%) |
| 17 | 4076 (9.1%) | 3991 (12.3%) | 8067 (10.4%) |
| 18 | 1674 (3.7%) | 2075 (6.4%) | 3749 (4.8%) |
| 19 | 942 (2.1%) | 1461 (4.5%) | 2403 (3.1%) |
| 20 | 496 (1.1%) | 850 (2.6%) | 1346 (1.7%) |
| **Gender** |  |  |  |
| Female | 23,589 (52.4%) | 16,500 (50.9%) | 40,089 (51.8%) |
| Male | 21,416 (47.6%) | 15,880 (49.0%) | 37,296 (48.2%) |
| **Ethnicity** |  |  |  |
| White | 16,446 (50.8%) | 17,964 (39.9%) | 34,410 (44.5%) |
| Black/AA | 6,019 (18.6%) | 12,012 (26.7%) | 18,031 (23.3%) |
| Hispanic | 4,925 (15.2%) | 9,629 (21.4%) | 14,554 (18.8%) |
| Other/Unknown | 4,995 (15.4%) | 5,402 (12.0%) | 10,397 (13.4%) |
| **Hospital** |  |  |  |
| A | 5,424 (12.1%) | 7,385 (22.8%) | 12,809 (16.6%) |
| B | 12,884 (28.6%) | 5,216 (16.1%) | 18,100 (23.4%) |
| C | 6,333 (14.1%) | 3,457 (10.7%) | 9,790 (12.6%) |
| D | 1,723 (3.8%) | 914 (2.8%) | 2,637 (3.4%) |
| E | 4,369 (9.7%) | 6,063 (18.7%) | 10,432 (13.5%) |
| F | 12,831 (28.5%) | 3,409 (10.5%) | 16,240 (21.0%) |
| G | 1,430 (3.2%) | 1,457 (4.5%) | 2,887 (3.7%) |
| H | 13 (0.0%) | 4,484 (13.8%) | 4,497 (5.8%) |
| **Entry time** |  |  |  |
| 07/2021 - 09/2021 | 38,335 (85.2%) | 24,509 (75.7%) | 62,844 (81.2%) |
| 10/2021 - 11/2021 | 6,672 (14.8%) | 7,876 (24.3%) | 14,548 (18.8%) |
| **Obesity** |  |  |  |
| 0 | 28,029 (62.3%) | 22,479 (69.4%) | 50,508 (65.3%) |
| 1 | 16,978 (37.7%) | 9,906 (30.6%) | 26,884 (34.7%) |
| **PMCA\*** |  |  |  |
| 0 | 25,634 (57.0%) | 19,916 (61.5%) | 45,550 (58.9%) |
| 1 | 10,915 (24.3%) | 6,417 (19.8%) | 17,332 (22.4%) |
| 2 | 8,458 (18.8%) | 6,052 (18.7%) | 14,510 (18.7%) |
| **Negative tests prior entry** |  |  |  |
| 0 | 1,330 (3.0%) | 2,888 (8.9%) | 4,218 (5.5%) |
| 1 | 34,272 (76.1%) | 16,299 (50.3%) | 50,571 (65.3%) |
| 2 | 7,388 (16.4%) | 9,739 (30.1%) | 17,127 (22.1%) |
| >=3 | 2,017 (4.5%) | 3,459 (10.7%) | 5,476 (7.1%) |

\*PMCA: Pediatric Medical Complexity Algorithm

**Table 2.** Baseline characteristics of children 5 to 11 and adolescents 12 to 20 years of age in the study of the effectiveness of the BNT162b2 vaccine in preventing infection and severe diseases with SARS-CoV-2 during periods when Omicron variant was prevalent.

|  | *Omicron study in children* |  |  | *Omicron study in adolescents* |  |  |
| --- | --- | --- | --- | --- | --- | --- |
|  | **Vaccinated**<br>**(N=50,398)** | **Unvaccinated**<br>**(N=61,141)** | **Overall**<br>**(N=111,539)** | **Vaccinated**<br>**(N=21,180)** | **Unvaccinated**<br>**(N=34,900)** | **Overall**<br>**(N=56,080)** |
| **Age** |  |  |  |  |  |  |
| **Median [Q1, Q3]** | 8 [6, 10] | 7 [6, 9] | 8 [6, 10] | 14 [13, 16] | 15 [13, 17] | 15 [13, 17] |
| **Distribution** |  |  |  |  |  |  |
| 5 | 8,165 (16.2%) | 13,321 (21.8%) | 21,486 (19.3%) |  |  |  |
| 6 | 7,447 (14.8%) | 11,314 (18.5%) | 18,761 (16.8%) |  |  |  |
| 7 | 7,090 (14.1%) | 9,151 (15.0%) | 16,241 (14.6%) |  |  |  |
| 8 | 7,028 (13.9%) | 7,922 (13.0%) | 14,950 (13.4%) |  |  |  |
| 9 | 6,773 (13.4%) | 7,085 (11.6%) | 13,858 (12.4%) |  |  |  |
| 10 | 7,011 (13.9%) | 6,434 (10.5%) | 13,445 (12.1%) |  |  |  |
| 11 | 6,884 (13.7%) | 5,914 (9.7%) | 12,798 (11.5%) |  |  |  |
| 12 |  |  |  | 4754 (22.4%) | 5760 (16.5%) | 10514 (18.7%) |
| 13 |  |  |  | 3421 (16.2%) | 5520 (15.8%) | 8941 (15.9%) |
| 14 |  |  |  | 3338 (15.8%) | 5299 (15.2%) | 8637 (15.4%) |
| 15 |  |  |  | 3123 (14.7%) | 5315 (15.2%) | 8438 (15.0%) |
| 16 |  |  |  | 2634 (12.4%) | 4944 (14.2%) | 7578 (13.5%) |
| 17 |  |  |  | 2201 (10.4%) | 3836 (11.0%) | 6037 (10.8%) |
| 18 |  |  |  | 986 (4.7%) | 2114 (6.1%) | 3100 (5.5%) |
| 19 |  |  |  | 475 (2.2%) | 1400 (4.0%) | 1875 (3.3%) |
| 20 |  |  |  | 248 (1.2%) | 712 (2.0%) | 960 (1.7%) |
| **Gender** |  |  |  |  |  |  |
| Female | 23,962 (47.5%) | 28,669 (46.9%) | 52,631 (47.2%) | 11,402 (53.8%) | 17,954 (51.4%) | 29,356 (52.3%) |
| Male | 26,436 (52.5%) | 32,468 (53.1%) | 58,904 (52.8%) | 9,775 (46.2%) | 16,939 (48.5%) | 26,714 (47.6%) |
| **Ethnicity** |  |  |  |  |  |  |
| White | 14,399 (28.6%) | 24,644 (40.3%) | 39,043 (35.0%) | 16,240 (46.5%) | 6,836 (32.3%) | 23,076 (41.1%) |
| Black/AA | 13,711 (27.2%) | 13,733 (22.5%) | 27,444 (24.6%) | 6,154 (17.6%) | 6,157 (29.1%) | 12,311 (22.0%) |
| Hispanic | 12,119 (24.0%) | 12,781 (20.9%) | 24,900 (22.3%) | 6,287 (18.0%) | 3,784 (17.9%) | 10,071 (18.0%) |
| Other/Unknown | 10,169 (20.2%) | 9,983 (16.3%) | 20,152 (18.1%) | 6,219 (17.8%) | 4,403 (20.8%) | 10,622 (18.9%) |
| **Hospital** |  |  |  |  |  |  |
| A | 5,019 (10.0%) | 9,266 (15.2%) | 14,285 (12.8%) | 2,131 (10.1%) | 5,183 (14.9%) | 7,314 (13.0%) |
| B | 15,229 (30.2%) | 13,168 (21.5%) | 28,397 (25.5%) | 6,397 (30.2%) | 6,556 (18.8%) | 12,953 (23.1%) |
| C | 5,482 (10.9%) | 7,409 (12.1%) | 12,891 (11.6%) | 1,719 (8.1%) | 4,075 (11.7%) | 5,794 (10.3%) |
| D | 4,766 (9.5%) | 2,878 (4.7%) | 7,644 (6.9%) | 678 (3.2%) | 1,337 (3.8%) | 2,015 (3.6%) |
| E | 5,843 (11.6%) | 10,551 (17.3%) | 16,394 (14.7%) | 2,047 (9.7%) | 4,263 (12.2%) | 6,310 (11.3%) |
| F | 9,786 (19.4%) | 11,348 (18.6%) | 21,134 (18.9%) | 3,563 (16.8%) | 5,186 (14.9%) | 8,749 (15.6%) |
| G | 1,250 (2.5%) | 3,239 (5.3%) | 4,489 (4.0%) | 622 (2.9%) | 2,353 (6.7%) | 2,975 (5.3%) |
| H | 3,023 (6.0%) | 3,282 (5.4%) | 6,305 (5.7%) | 4,023 (19.0%) | 5,947 (17.0%) | 9,970 (17.8%) |
| **Entry time** |  |  |  |  |  |  |
| 01/2022 - 03/2022 | 37,970 (75.3%) | 32,523 (53.2%) | 70,493 (63.2%) | 14,684 (69.3%) | 1,9032 (54.5%) | 33,716 (60.1%) |
| 04/2022 - 06/2022 | 5,882 (11.7%) | 11,919 (19.5%) | 17,801 (16.0%) | 3,344 (15.8%) | 7,087 (20.3%) | 10,431 (18.6%) |
| 07/2022 - 09/2022 | 4,994 (9.9%) | 10,329 (16.9%) | 15,323 (13.7%) | 2,206 (10.4%) | 5,479 (15.7%) | 7,685 (13.7%) |
| 10/2022 - 11/2022 | 1,552 (3.1%) | 6,370 (10.4%) | 7,922 (7.1%) | 946 (4.5%) | 3,302 (9.5%) | 4,248 (7.6%) |
| **Obesity** |  |  |  |  |  |  |
| 0 | 33,381 (66.2%) | 42,165 (69.0%) | 75,546 (67.7%) | 13,832 (65.3%) | 23,895 (68.5%) | 37,727 (67.3%) |
| 1 | 17,017 (33.8%) | 18,976 (31.0%) | 35,993 (32.3%) | 7,348 (34.7%) | 11,005 (31.5%) | 18,353 (32.7%) |
| **PMCA\*** |  |  |  |  |  |  |
| 0 | 33,870 (67.2%) | 40,976 (67.0%) | 74,846 (67.1%) | 13,482 (63.7%) | 21,079 (60.4%) | 34,561 (61.6%) |
| 1 | 10,000 (19.8%) | 11,189 (18.3%) | 21,189 (19.0%) | 4,382 (20.7%) | 6,764 (19.4%) | 11,146 (19.9%) |
| 2 | 6,528 (13.0%) | 8,976 (14.7%) | 15,504 (13.9%) | 3,316 (15.7%) | 7,057 (20.2%) | 10,373 (18.5%) |
| **Negative tests prior entry** |  |  |  |  |  |  |
| 0 | 2,337 (4.6%) | 5,640 (9.2%) | 7,977 (7.2%) | 768 (3.6%) | 2,966 (8.5%) | 3,734 (6.7%) |
| 1 | 34,077 (67.6%) | 28,417 (46.5%) | 62,494 (56.0%) | 15,707 (74.2%) | 17,303 (49.6%) | 33,010 (58.9%) |
| 2 | 10,514 (20.9%) | 19,816 (32.4%) | 30,330 (27.2%) | 3,654 (17.3%) | 11,012 (31.6%) | 14,666 (26.2%) |
| >=3 | 3,470 (6.9%) | 7,268 (11.9%) | 10,738 (9.6%) | 1,051 (5.0%) | 3,619 (10.4%) | 4,670 (8.3%) |

\*PMCA: Pediatric Medical Complexity Algorithm

### VACCINE EFFECTIVENESS

Table 3 summarizes the estimated vaccine effectiveness in three study cohorts and Figure 2 shows the durability of protection. The vaccine effectiveness was estimated to be 98.4% (95% CI, 98.1 to 98.7) among adolescents in the Delta period, 74.3% (95% CI, 72.2 to 76.2) against documented infection among children in the Omicron period, and 85.5% (95% CI, 83.8 to 87.1) among adolescents in the Omicron period. During the Delta period, the vaccine effectiveness against documented infection remained stable throughout the follow-up period of the study. After 4 months following the first dose, vaccine effectiveness against documented infection with Omicron declined from 82.3% (95% CI, 77.9 to 85.8) to 70.6% (95% CI, 65.9 to 74.6) among children, and from 91.3% (95% CI, 87.6 to 94.0) to 82.9% (95% CI, 79.0 to 86.1) among adolescents. Although vaccine effectiveness against documented infection stabilized after this initial decline, the corresponding confidence intervals were much wider indicating higher levels of uncertainty.

**Figure 1.**
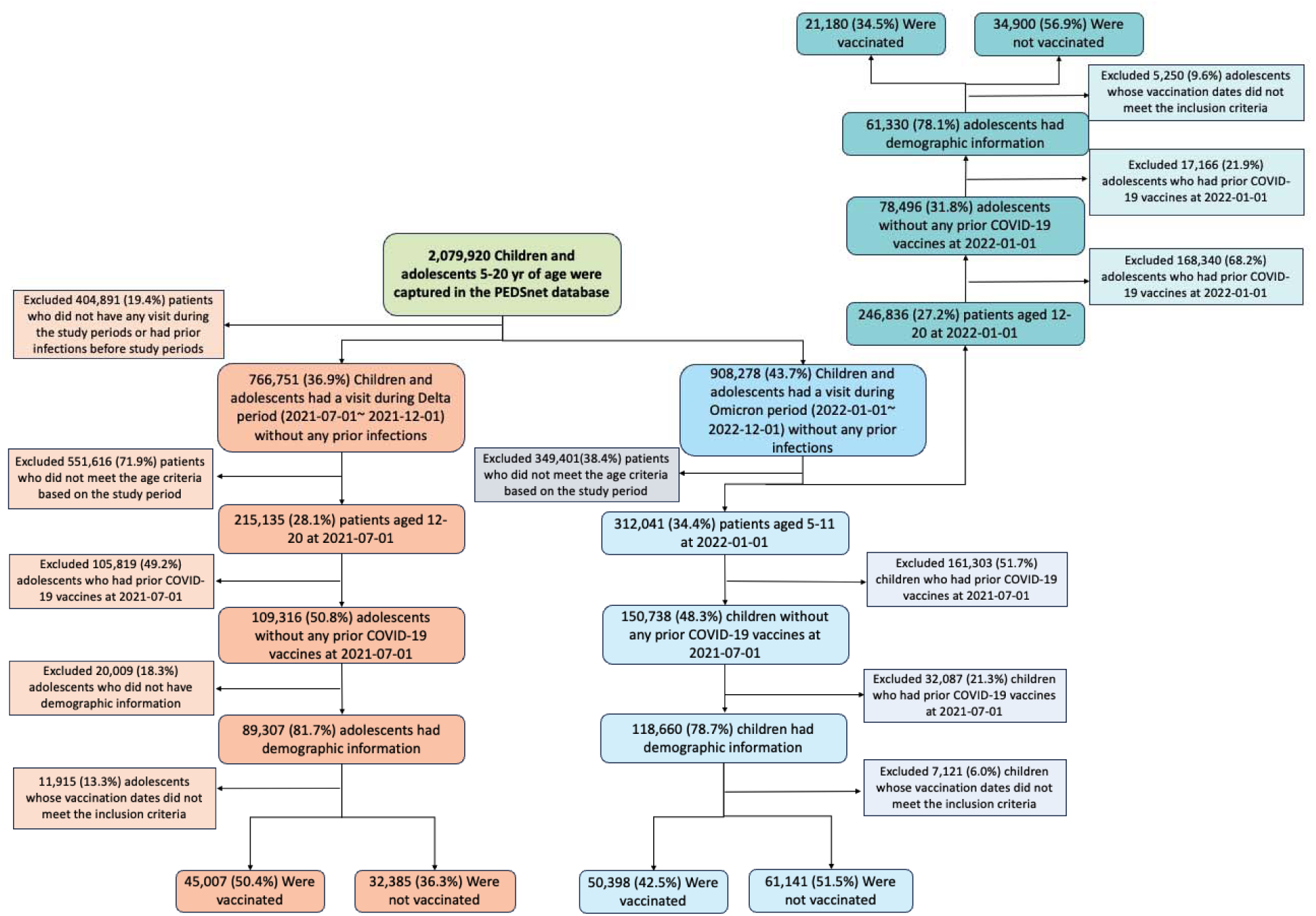
Selection of participants for the three study cohorts evaluating the effectiveness of the BNT162b2 vaccine in preventing infection with SARS-CoV-2 in (*Study cohort 1*) adolescents aged 12-20 years during the period when the Delta variant was prevalent, (*Study cohort 2*) children aged 5 to 11 years and (*Study cohort 3*) adolescents aged 12 to 20 years during the period when the Omicron variant was prevalent.

**Figure 2.**
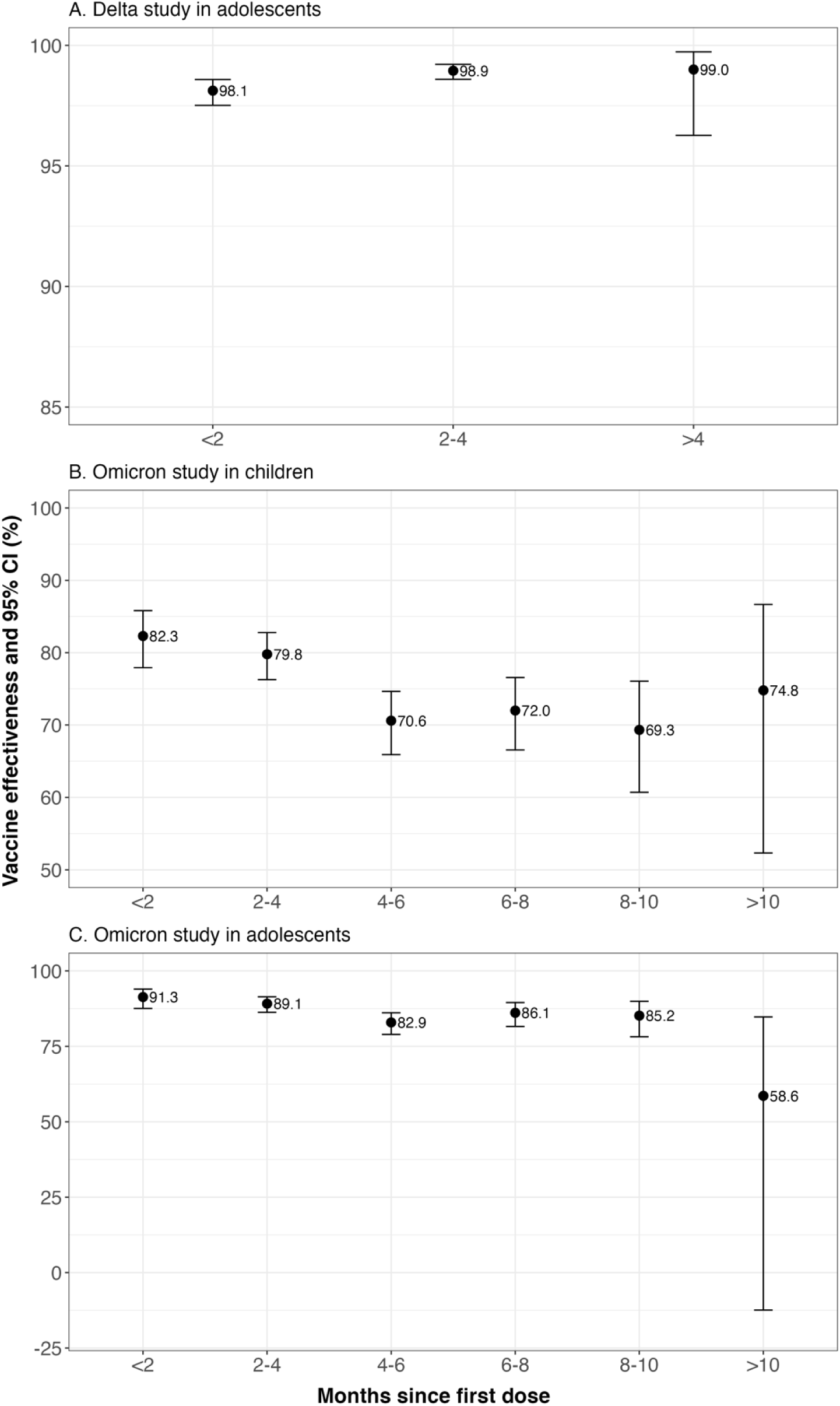
Stratified effectiveness of the BNT162b2 vaccine in preventing infection with SARS-CoV-2 in children and adolescents by 2-month intervals since receipt of the first dose

**Table 3.** Estimated effectiveness of the BNT162b2 vaccine in preventing infection and severe.

|  | Vaccinated | Unvaccinated | Overall | Vaccine effectiveness (in %) and 95% CI |
| --- | --- | --- | --- | --- |
| *Delta study in adolescents* |  |  |  |  |
| **Follow-up** |  |  |  |  |
| Total follow-up — no. of person-wk | 644,162 | 398,906 | 1,043,068 |  |
| Median [Q1, Q3] | 16 [12, 18] | 13 [9, 17] | 15 [10, 18] |  |
| **Absolute risk (in %)** |  |  |  |  |
| Documented infection | 0.35 | 5.26 | 2.41 | 98.4 (98.1, 98.7) |
| Mild COVID-19 | 0.06 | 1.43 | 0.63 | 99.0 (98.5, 99.3) |
| Moderate or severe COVID-19 | 0.03 | 0.49 | 0.22 | 98.7 (97.4, 99.3) |
| ICU* admission with COVID-19 | <0.01 | 0.05 | < 0.03 | 99.0 (92.5, 99.9) |
| Cardiac complication | 0.02 | 0.03 | 0.02 | 1.22 (0.34, 4.35)<sup>†</sup> |
| **Age 12-15 yr** |  |  |  |  |
| Total follow-up — no. of person-wk | 458,981 | 229,083 | 688,064 |  |
| Documented infection | 0.34 | 5.52 | 2.28 | 99.0 (98.6, 99.3) |
| **Age 16-21 yr** |  |  |  |  |
| Total follow-up — no. of person-wk | 185,181 | 169,823 | 355,003 |  |
| Documented infection | 0.36 | 4.91 | 2.64 | 97.0 (95.9, 97.8) |
| *Omicron study in children* |  |  |  |  |
| **Follow-up** |  |  |  |  |
| Total follow-up — no. of person-wk | 1,925,686 | 1,911,599 | 3,837,285 |  |
| Median [Q1, Q3] | 44 [35, 46] | 36 [19, 44] | 40 [25, 45] |  |
| **Absolute risk (in %)** |  |  |  |  |
| Documented infection | 1.89 | 5.46 | 3.85 | 74.3 (72.2, 76.2) |
| Mild COVID-19 | 0.54 | 1.55 | 1.09 | 73.5 (69.2, 77.1) |
| Moderate or severe COVID-19 | 0.19 | 0.67 | 0.45 | 75.5 (69.0, 81.0) |
| ICU admission with COVID-19 | 0.02 | 0.08 | 0.05 | 84.9 (64.8, 93.5) |
| Cardiac complication | <0.01 | 0.03 | <0.02 | 0.28 (0.08, 0.95)<sup>†</sup> |
| **Age 5-8 yr** |  |  |  |  |
| Total follow-up — no. of person-wk | 1,101,418 | 1,254,819 | 2,356,236 |  |
| Documented infection | 1.96 | 5.10 | 3.78 | 71.3 (68.2, 74.1) |
| **Age 9-11 yr** |  |  |  |  |
| Total follow-up — no. of person-wk | 824,268 | 656,780 | 1,481,049 |  |
| Documented infection | 1.80 | 6.11 | 3.95 | 77.9 (75.1, 80.4) |

| *Omicron study in adolescents* |  |  |  |  |
| --- | --- | --- | --- | --- |
| **Follow-up** |  |  |  |  |
| Total follow-up — no. of person-wk | 772,176 | 1,113,561 | 1,885,736 |  |
| Median [Q1, Q3] | 42 [30, 45] | 37 [22, 44] | 39 [25, 45] |  |
| **Absolute risk (in %)** |  |  |  |  |
| Documented infection | 1.82 | 8.17 | 5.77 | 85.5 (83.8, 87.1) |
| Mild COVID-19 | 0.43 | 2.07 | 1.45 | 87.0 (83.5, 89.8) |
| Moderate or severe COVID-19 | 0.15 | 0.88 | 0.60 | 84.8 (77.3, 89.9) |
| ICU admission with COVID-19 | <0.02 | 0.14 | < 0.10 | 91.5 (69.5, 97.6) |
| Cardiac complication | <0.02 | 0.05 | <0.04 | 0.10 (0.02, 0.57)<sup>†</sup> |
| **Age 12-15 yr** |  |  |  |  |
| Total follow-up — no. of person-wk | 524,053 | 654,315 | 1,178,368 |  |
| Documented infection | 1.93 | 8.24 | 5.67 | 85.8 (83.6, 87.7) |
| **Age 16-21 yr** |  |  |  |  |
| Total follow-up — no. of person-wk | 248,123 | 459,246 | 707,368 |  |
| Documented infection | 1.60 | 8.07 | 5.93 | 85.9 (82.7, 88.5) |

\*: ICU: Intensive Care Unit
†: relative risk in the vaccinated groups compared to the unvaccinated.

### VACCINE EFFECTIVENESS: SEVERE ILLNESS AND COMPLICATIONS

During the Delta period, the vaccine was found to have high effectiveness against severe infections. The estimated relative risk the vaccine on cardiac complications was 1.22 (95% CI, 0.34 to 4.35). The estimated vaccine effectiveness against the Omicron variant in children was 73.5% (95% CI, 69.2 to 77.1) against mild COVID-19, 75.5% (95% CI, 69.0 to 81.0) against moderate or severe COVID-19, and 84.9% (95% CI, 64.8 to 93.5) against ICU admission with COVID-19. The estimated relative risk the vaccine on cardiac complications was 0.28 (95% CI, 0.08 to 0.95). In the Omicron study in adolescents, the vaccine effectiveness was estimated to be 87.0% (95% CI, 83.5 to 89.8) against mild COVID-19, 84.8% (95% CI, 77.3 to 89.9) against moderate or severe COVID-19, and 91.5% (95% CI, 69.5 to 97.6) against ICU admission with COVID-19. The estimated relative risk of the vaccine on cardiac complications was 0.10 (95% CI, 0.02 to 0.57) in this cohort.

### SENSITIVITY ANALYSES AND ADDRESSING MISCLASSIFICATION BIAS

Section S9 presents the vaccine effectiveness against various Omicron sub-variants. The effectiveness against sub-variants BA.1, BA.2, BA.4, and BA.5 aligns with our primary findings, while the vaccine’s effectiveness appears to be lower against the BQ.1, XBB, and subsequent sub-variants. Section S14 presents negative control experiments of three study cohorts using 40 negative control outcomes. After accounting for systematic error through calibration using negative control outcomes, our findings indicate a slight shift in point estimates accompanied by wider confidence intervals. This suggests the presence of a minor degree of systematic error, as well as additional uncertainty characterized by the estimated distribution derived from the negative control outcomes. Section S19 summarizes the comparative results to a sequential target trial emulation not accounting for underreporting issues of vaccination, which indicated reasonably consistent findings.

Section S16 shows effectiveness estimated from the naive method and proposed CER method with different ranges of sensitivity of vaccination status captured by EHR. The comparison results indicated that the vaccine effectiveness was reasonably consistent across different sensitivity ranges, suggesting that our primary analysis was robust to changes in the range of sensitivity considered. Section S17 shows the comparative results to a fully Bayesian method indicating nearly identical results. Section S18 shows sensitivity analyses on differential misclassifications which demonstrates the novel CER method corrects the bias even when the non-differential misclassification assumption does not hold.

## DISCUSSION

We estimated the effectiveness of BNT162b2 vaccines for the prevention of documented COVID-19 infections and severe disease in a national network of pediatric health systems in the U.S. for three study cohorts. During the period of time where the Delta variant was dominant, the BNT162b2 vaccine in adolescents was associated with strong protection with effectiveness higher than 95% and with little evidence of waning during the follow-up period. Our findings against the Delta infection among adolescents are consistent with vaccine efficacy observed in the BNT162b2 clinical trial involving adolescents between 12 and 15 years of age, which demonstrated vaccine efficacy of 100% (95% CI, 75.3 to 100) against confirmed COVID-19 infection(2). Our estimates of vaccine effectiveness against severe diseases are consistent with a case-control study based on test-negative design, which found an effectiveness of 94% (95% CI, 90 to 96) against hospitalization and 98% (95% CI, 93 to 99) against ICU admission(7).

In our study, during the period of time where the Omicron variant was dominant, the estimated vaccine effectiveness was approximately 70% for children and 85% for adolescents. The estimated protection decreased by roughly 10% around four months from the first dose and slightly waned over time. Previous studies have shown vaccine effectiveness against Omicron infection, ranging from 20 to 80% among children and adolescents. An analysis using a test-negative design in Scotland during Omicron period revealed a vaccine effectiveness of 81.2% (95% CI, 77.7 to 84.2) for adolescents aged 12 to 15 years, and 65.5% (95% CI, 56.0 to 73.0) for those aged 16 to 17 years, 2-5 weeks post full vaccination. This effectiveness decreased to 43.3% (95% CI, 30.0 to 54.2) and 8.9% (95% CI, –19.1 to 30.3) after 10-13 weeks, respectively(20). With a test-negative design, data from U.S. pharmacy-based, drive-through SARS-CoV-2 testing sites indicated that the estimated vaccine effectiveness for children aged 5 to 11 years was 60.1% within 2 to 4 weeks following the second dose and declined to 28.9% during the second month post-vaccination. For adolescents aged 12 to 15 years, the effectiveness was 59.5% within 2 to 4 weeks after the second dose and dropped to 16.6% during the subsequent month (21). Another test-negative design analysis in U.S. indicated that in children aged 5–11 years, the effectiveness was 59.9% (95% CI 58.5 to 61.2) at 1 month, 33.7% (32.6 to 34.8) at 4 months, and 14.9% (95% CI 12.3 to 17.5) at 10 months following the first dose(22). A retrospective study among Italy children aged 5-to-11 years shown the vaccine effectiveness to be 29.4% (95% CI 28.5 to 30.2) against documented infection and 41.1% (95% CI 22.2 to 55.4) against severe COVID-19(23). A Singapore study founded that in fully vaccinated children aged 5-to-11 years, vaccine effectiveness was 65.3% (95% CI, 62.0 to 68.3) against PCR-confirmed SARS-CoV-2 infection and 82.7% (95% CI, 74.8 to 88.2) against COVID-19-related hospitalization (24). Additionally, we found that the vaccine effectiveness is substantially lower against the later sub-variants of Omicron including BQ.1, XBB. However, it remains unclear whether this is a true sub-variant effect or evidence of further waning over time. Continued research is desirable to understand the vaccine effectiveness on future sub-variants and its potential waning effects.

This study did not identify a statistically significant elevated risk of cardiac complications among vaccinated adolescents during the Delta variant period, and it even demonstrated a lower risk in the vaccinated group during the Omicron variant period. This finding might seem unexpected, given that cases of myocarditis and pericarditis after mRNA COVID-19 vaccines received significant attention(25–27). However, it is essential to note that previous studies indicated a much higher risk of myocarditis or pericarditis following a documented COVID-19 infection in the pediatric population(28), with one paper finding 36.8 times higher risk (95% CI 25.0 to 48.6) in less than 16 years old after COVID-19(29), and that myocarditis is a common symptom for patients diagnosed with MIS(30,31). It is possible cases of myocarditis, pericarditis or MIS occurred after undocumented COVID-19 infections. For example, 31.5% of occurrences of MIS did not have COVID-19 infections documented in the PEDSnet EHR database. Also note that during this period, especially the Omicron period, positive tests may have been from at-home tests or otherwise outside the system. Further assessment of cardiac complications after vaccination and COVID-19 is warranted to help provide a more complete picture of risk or benefit during a changing pandemic.

Our study has several strengths. First, we used a national network of academic medical centers that covered a diverse cohort being more representative of the general pediatric population, provided a robust sample size, and allowed for multiple subgroup analyses and detection of rare outcomes. Second, the richness of these EHR data allowed us to investigate the effectiveness against infection of different levels of severity as well as adjust for a broad set of confounders. Third, we conducted the negative control outcome experiments to assess the potential residual bias due to unmeasured confounders and other potential sources of systematic bias in the data. These experiments revealed a small amount of systematic error but with excessive uncertainty across different negative control outcomes, leading to wider confidence intervals of our estimated effectiveness that honestly reflect the impacts of unmeasured confounding and other potential sources of residual biases(32). Finally, to the best of our knowledge, this is the first real-world effectiveness study evaluating COVID-19 vaccines against infection and severe outcomes that explicitly handle the underreporting in vaccination.

Our study also has several potential limitations. First, effectiveness was investigated against documented infection in a cohort without previous infection, while the potential inclusion of patients with undocumented infections exists. Nevertheless, if this potential lack of data is evenly distributed across treatment arms, it could potentially attenuate the true vaccine effectiveness, thereby making our analysis more conservative in its estimations. Our inclusion of previous negative COVID-19 tests as a confounder aims to balance the probability of testing between treatment arms which could partially adjust for this factor. Moreover, the increasing availability of at-home rapid antigen testing kits over time could have further reduced the testing frequency captured by EHR. However, severe cases who test positive through home kits typically seek medical care and report their results to hospitals. This would lead to a better capture of severe infection in our database and more reliable vaccine effectiveness estimates. Baseline confounders were balanced between vaccinated and unvaccinated groups, which should adjust for between-cohort differences in exposure risks and risks of severe infections. Second, as in any observational study, assignment to the vaccine group was non-random and the validity of the results could be impacted by unmeasured confounders. To evaluate the impact of unmeasured confounders and residual bias, we conducted negative control experiments that quantified the robustness of our results.

Third, in this study, patients who had received vaccinations prior to the start of the study period were excluded. Due to missing vaccine records, some patients who had previously been vaccinated may have still entered the cohort, particularly in the unvaccinated group. However, the CER method used in this study adjusted for potential bias resulting from unrecorded vaccinated patients which could also reduce the bias resulting from this issue. Finally, in the Omicron study involving adolescents, the cohort included adolescents who had their first vaccine after January 1, 2022. Since the use of BNT162b2 vaccines was authorized in adolescents aged 12-15 years on May 10, 2021, this cohort may represent a population with late vaccines which reduces the generalizability of the findings.

Although this study provides evidence of a slight waning of vaccine effectiveness 4 months following the first dose against Omicron infection and the effectiveness is stabilized after 4 months, waning can be impacted by vaccines during the follow-up period and other factors. Patients who got boosters during the follow-up period were not excluded from the study. A sensitivity analysis evaluating the durability of two-dose vaccine effectiveness considering the third dose as censoring did not suggest a significantly different conclusion. A future study is warranted to investigate the effect of booster vaccination among children and adolescents. Furthermore, despite the recognized risk of myocarditis associated with COVID-19 vaccines in young men and teen boys, the study reveals a lower relative risk of myocarditis, pericarditis, or MIS in vaccinated groups which may be partially explained by reduced likelihood of infection during the study period(33).

In summary, this study involving national pediatric cohorts in the U.S. estimates moderate effectiveness of the BNT162b2 vaccine for preventing infection and severe diseases of the SARS-CoV-2 Omicron variant, and high effectiveness against the Delta variant. This study reveals a low risk of cardiac complications among children and adolescents who were vaccinated during the Omicron period, which was statistically insignificant in the Delta period. Our assessment of vaccine effectiveness across diverse outcomes underscores the vaccine’s pivotal role in reducing SARS-CoV-2 transmission, minimizing COVID-19 related sick leaves, and alleviating economic burdens during the pandemic. This study significantly contributes to our knowledge of the BNT162b2 vaccine in the U.S. pediatric population using a rigorously designed CER method accounting for the incomplete capture of vaccination status in EHR data in the U.S.

## Data Availability

All data produced in the present study are available upon reasonable request to the authors

